# Improving the care of children with GENetic Rare disease: Observational Cohort study (GenROC): a study protocol

**DOI:** 10.1101/2024.02.23.24303131

**Authors:** KJ Low, A Watford, PS Blair, I Nabney, J Powell, SL Wynn, J Foreman, HV Firth, JC. Ingram

**Affiliations:** Centre for Academic Child Health, Bristol Medical School, University of Bristol; Department of Clinical Genetics, University Hospitals Bristol and Weston NHS Trust, Bristol, UK; School of Computer Science, Electrical and Electronic Engineering and Engineering Maths, University of Bristol; Nuffield Department of Primary Care Health Sciences, University of Oxford; Unique, Rare Chromosome Disorder Support Group, Oxted, UK; European Molecular Biology Laboratory, European Bioinformatics Institute, Wellcome Genome Campus, Hinxton, Cambridge, CB10 1SD, United Kingdom; Department of Clinical Genetics, Addenbrookes hospital, Cambridge, UK; Wellcome Sanger Institute, Wellcome Genome Campus, Hinxton, Cambridge, CB10 1SA

**Keywords:** Clinical Genetics and dysmorphology, Paediatrics, Paediatric neurology, Community child health

## Abstract

Around 2000 children are born in the UK per year with a neurodevelopmental genetic syndrome with significantly increased morbidity and mortality(1). Often little is known about expected growth and phenotypes in these children. Parents have responded by setting up social media groups to generate data themselves. Given the significant clinical evidence gaps, this research will attempt to identify growth patterns, developmental profiles and phenotypes, providing data on long-term medical and educational outcomes. This will guide clinicians when to investigate, monitor or treat symptoms and when to search for additional or alternative diagnoses.

**Methods and analysis:** This is an observational, multicentre cohort study recruiting between March 2023 and February 2026. Children aged 6 months up to 16 years with a pathogenic or likely pathogenic variant in a specified gene will be eligible. Children will be identified through the NHS and via self-recruitment. Parents or carers will complete a questionnaire at baseline and again one year after recruitment. The named clinician (in most cases a clinical geneticist) will complete a clinical proforma which will provide data from their most recent clinical assessment. Qualitative interviews will be undertaken with a subset of parents partway through the study. Growth and developmental milestone curves will be generated through the DECIPHER website (https://deciphergenomics.org) where 5 or more children have the same genetic syndrome (at least ten groups expected).

The results will be presented at national and international conferences concerning the care of children with genetic syndromes. Results will also be submitted for peer review and publication.

**Article Summary:** Strengths and Limitations of this study

- This study is a collaborative effort which combines previous data with that of a large cohort to maximise possible outputs through the DECIPHER database.
- The study is the first of its kind to acquire natural history clinical data from a large cohort across the UK to ascertain both gene specific data but also cross syndrome cohort metrics.
- The study utilises the Musketeers Memorandum to facilitate recruitment from genetics centres across the UK and also empowers families to volunteer directly which will be supported by a national patient support charity.
- This study does not recruit individuals over the age of 15 and so longer-term outcomes will not be evaluated. Further studies of older individuals will be required to gather their data and we hope that a follow-on study would be possible to enable this.
- Data are largely gathered through online questionnaires completed by both parent and clinician. The quality of data received will vary depending on the user. This method of data collection may result in selection bias against certain demographic groups which will need to be accounted for in any discussion regarding the generalisability of the results.

## Introduction

Genetic syndromes are a major contributor to demands on paediatric healthcare services. There are almost 2000 different genes currently known to be associated with neurodevelopmental syndromes with an estimated 2000 children in the UK diagnosed each year(2). Genetic disorders are found in 20-30% of cases of all infant deaths. An estimated 12% of paediatric admissions are related to congenital anomalies and genetic disorders, with a mortality 4.5 times greater than other children who had been admitted. Many of these deaths are within the first year of life(3, 4).

For most syndromes there is some information on the associated dysmorphic features and medical complications, but data are limited regarding natural history, range of growth patterns and range of clinical phenotypes. This prevents clinicians knowing whether a clinical feature is expected or requires further investigation, monitoring or treatment. There is wide variation between syndromes with respect to availability of sufficient resources for health professionals or families. Rare syndromes collectively represent a substantial disease burden(1). The GenROC study aims to improve the understanding of natural history phenotypes in this group of rare syndromes in order to better inform clinical decision making.

## Methods and analysis

### Study design

GENROC is a prospective UK cohort study of 500 children with genetic syndromes.

Eligibility criteria are as follows: Individual with a clinical diagnosis of intellectual disability/ developmental delay; previously identified pathogenic or likely pathogenic mutation in one of the specified genes (see gene list supplementary figure); and aged 6 months-15 years. The following conditions would render the child ineligible: Genetic mutation classed as benign/likely benign or of uncertain significance; Mutation in a gene not specified in the list; Chromosomal disorders; diagnosis identified by Deciphering Developmental Disorders (DDD) Study; Composite genetic diagnosis(where participant has more than one genetic diagnosis that is contributing to their phenotype as assessed by their clinician).

The study is designed to maximise phenotypic data collection through multiple sources, namely from parents, clinicians and from digital sources such as the internet (gene specific sites), social media (such as Facebook groups). This multi-pronged strategy is depicted in the GENROC flowchart (figure1).

**Figure 1:**
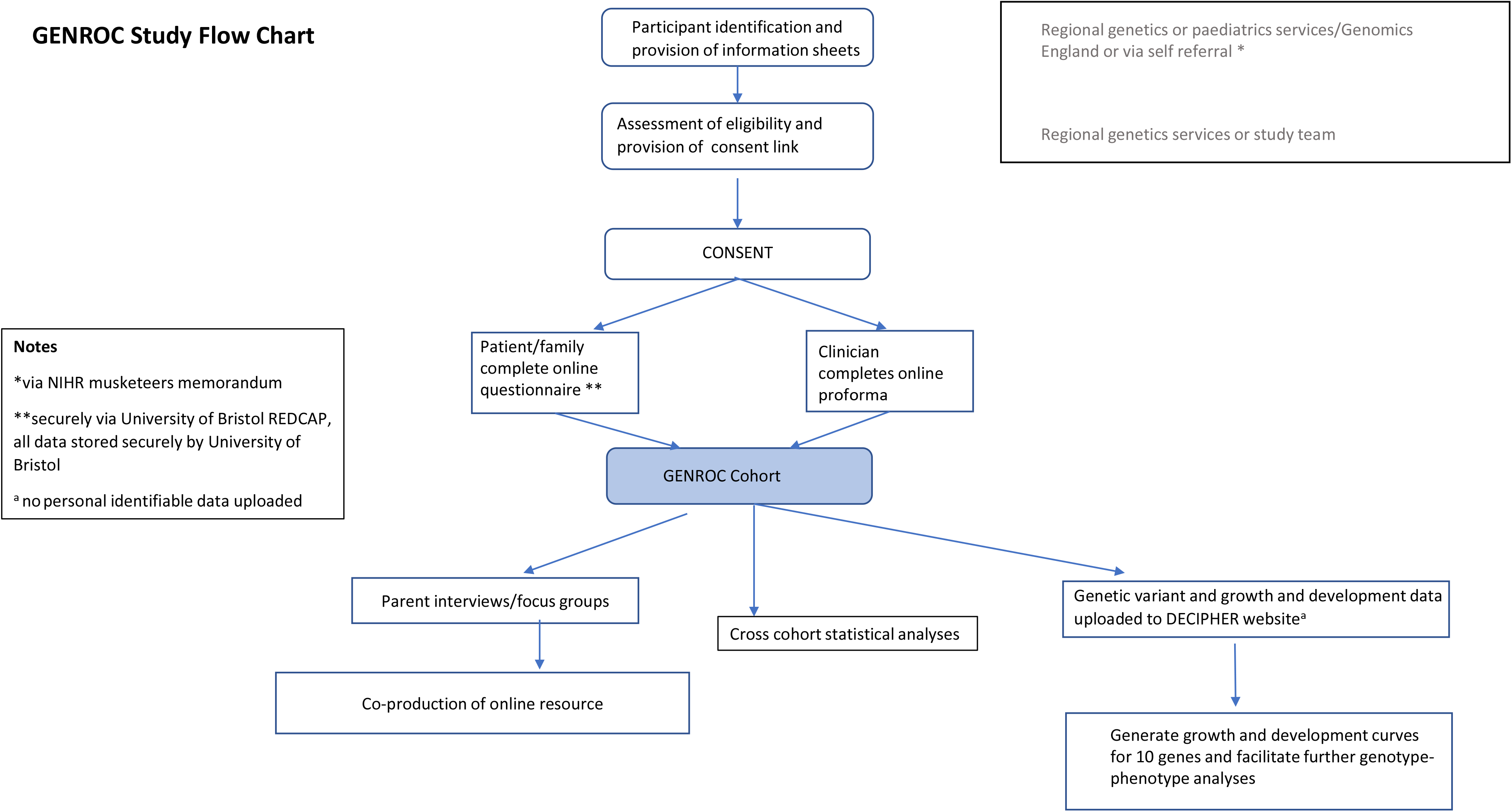
GenROC Study Flow Chart.

Children will be recruited from NHS clinical genetics centres through the Musketeers Memorandum(5). There are 23 clinical genetics centres which deliver paediatric genetics clinics across the UK. Children with genetic syndromes are tested and seen by paediatricians in multiple other settings such as community paediatrics and tertiary hospital settings. Children can be recruited through these paediatricians as an additional avenue of recruitment. Families will also be able to refer themselves directly should they become aware of the study through publicity.

It may also become possible for families to be contacted about the study via Genomics England. These would be children who were diagnosed through the 100,000 Genomes project(6) and whose parents have consented to being contacted about further research studies.

The DECIPHER project (www.deciphergenomics.org) enables clinicians and scientists around the world to share information about rare genomic variants and the patient’s clinical symptoms to facilitate diagnosis of known disorders and to help to elucidate the role of genes whose function is not yet known(7). The interface for recording morphometry, milestones and pregnancy history was added around 2011 when the Deciphering Developmental Disorders Study(2) started as an initiative to populate DECIPHER with data from consistently phenotyped and genotyped patients. In subsequent years, the interfaces which display the models for individual genetic disorders have been developed and now show how an individual patient fits with the phenotype-gene-disease model and how good a diagnostic fit a candidate diagnosis provides. The DECIPHER development team have designed a computer-based growth chart tool which can produce gene specific growth curves and/or developmental milestone ranges once head circumference, height, weight or milestone data from a minimum of 5 children with a pathogenic/likely pathogenic variant in that gene has been input. The DECIPHER platform depicts this growth curve alongside a normal paediatric growth curve. The platform also shows the clinical features found in diagnosed patients in descending order of frequency and the most discriminatory features. With increased data, the utility of the growth tools will be greatly enhanced.

### Sample size

We plan to recruit 500 participants. This pragmatic sample size was based on the estimate of 2000 children being diagnosed with a genetic rare disease each year in the UK(2) and on the basis of a high number of relatively new diagnoses that have been made in the last 5 years due to the DDD study, the 100,000 genomes project and the introduction of rapid exome sequencing and whole genome sequencing into the NHS. We have calculated that this sample size would enable us to recruit at least 5 participants for at least ten different genes enabling production of gene specific growth curves on DECIPHER. Given the need for at least 5 patients per syndrome to generate DECIPHER curves we have curated a list of genes to maximise the chance of recruiting the minimum threshold number of participants for as many genes as possible within the total cohort of 500. This has been determined based on the strategy employed by Aitken et al with IMPROVE-DD(8). The gene list generated by Aitken et al has been reviewed and genes have been excluded where a) they are already being studied in the UK as part of a large phenotypic study or b) where growth curves already exist. The Gene2Phenotype Developmental Disorders panel gene list (9)has then been reviewed on DECIPHER and filtered according to syndromes associated with OMIM Morbid (Home -OMIM) (10) intellectual disability/developmental delay syndrome and then by numbers of patients. This strategy resulted in the final gene list of 125 genes (see supplementary figure 1) which represents the most frequently diagnosed causes of rare developmental disorders from the DDD study and DECIPHER. Analysis of 100,000 Genomes confirmed diagnoses for eligibility has determined approximately 500 potentially eligible participants.

The DECIPHER automated curves require a minimum dataset of 5 individuals per gene. We therefore estimate that our sample size of 500 will allow us to recruit the minimum dataset for at least ten of the listed genes. DECIPHER already holds data for many of these genes and so even for the genes for which we are not able to recruit 5 it is likely that we will be adding to the existing dataset which will enable curves to be produced by combining data.

### Data collection

Observations will be obtained at two time points for each participant (Table1). The first time point will be at the age of last clinical assessment through retrospective review of clinical records. The second time point will be at the time of recruitment via parent reported questionnaire. Following consent, the child’s named clinician (in most cases their clinical geneticist) will be sent an online secure link to a clinical proforma in Research Electronic Data Capture (REDCap) a web-based data capture system for research. They will be required to complete a minimum data set including genomic variant information, last set of anthropomorphic measurements (height, weight, head circumference) and HPO Terms to describe presence of phenotypic features consistently (11). The clinical questionnaire has been derived from the Deciphering Developmental Disorders (DDD) Study phenotyping questionnaire and DECIPHER(7), converted into lay terminology and then combined with several measures studied through other cohort studies such as IMAGINE-ID(12). The parent questionnaire was co-produced with PPI members and subsequently user tested by 3 parents and revised based on their feedback. Parents will be asked to complete a short follow up questionnaire 12-18 months after recruitment (to allow for the phased opening of sites and staggered recruitment initially).

**Table 1:** Data collection schedule.

| Data Item |  | Baseline | Time point1 | Timepoint 2 |
| --- | --- | --- | --- | --- |
| Assessment data | Name, date of birth, address, NHS number | x |  |  |
|  | Sex | x |  |  |
|  | Ethnicity | x |  |  |
|  | Genetic mutation, classification and test methodology | x |  |  |
|  | Clinical phenotypes | x |  |  |
|  | Developmental history | x |  |  |
|  | Pedigree | x |  |  |
|  | Pregnancy history | x |  |  |
|  | Birth and last documented height, weight and head circumference | x |  |  |
| Questionnaire completed by parent/carer | Developmental and pregnancy history | x |  |  |
|  | Medical problems | x |  | x |
|  | Behaviour problems | x |  | x |
|  | Current height, weight and head circumference | x |  | x |
|  | Family history | x |  | x |
|  | Current education setting, level of support/assessment and function | x |  | x |
|  | Parental participation in social media groups | x |  | x |
|  | Parental use of digital healthcare tools | x |  | x |
|  | Healthcare utilisation and experience | x |  | x |
| Qualitative focus group/interviews | Parental experience of diagnosis |  | x |  |
|  | Parental views/experience of data availability |  | x |  |
|  | Participation in and Data sharing in social media groups |  | x |  |
|  | Views/concerns around identifiable data |  | x |  |
|  | Parental views on possible outputs from study |  | x |  |

We will collect qualitative data through parent interviews. Parents will have had an option to consent to interviews at the end of their questionnaire and we will select parents purposively for a range of characteristics by genetic diagnosis, postcode and gender. We will select parents based on questionnaire answers aiming to include parents who don’t use social media as well as those who are very active in these groups. Topic guides will be informed by the research literature, team discussions and input from PPI and will include questions around the diagnostic journey, parental experience and attitudes to information about their child’s condition and about their utilisation of social media and the internet in this regard. Selected parents will be invited by email and provided with further information. We will conduct semi-structured interviews either face to face, online or on the phone. It is anticipated interviews will last between 30min and 1 hour.

## Outcome measures and data analysis plan

### Quantitative data analysis

DECIPHER tools already exist to generate growth curves for genes. Growth curves will then be automatically generated by DECIPHER following upload of the data of sufficient participants per gene. GENROC data will also be used to generate other gene specific DECIPHER automated tools such as lists of common phenotypic features and developmental trajectory curves.

A STROBE flow diagram will be produced which will include numbers of individuals at each stage of study—numbers potentially eligible, confirmed eligible, included in the study, completing follow-up, and analysed. A statistical analysis plan will be written before the final participant is recruited and prior to data analysis. The plan will be available on request. Demographic, clinical, social characteristics of study participants will be summarised with descriptive statistics. Means or medians together with appropriate measures of dispersion will be reported for continuous measures and proportions for binary measures. The study will use standard inference statistical methods to compare clinical features of study participants with general population data. For analyses involving a larger number of variables, statistical methods adapted to such datasets will be employed. For the top ten conditions gene specific analyses will be undertaken and statistical methods adapted accordingly. comparisons will be stated using a mixed methods approach which may include topic and sentiment modelling and natural language processing applied to the datasets. Any subsequent exploratory analyses will be labelled as such when reported.

### Qualitative data analysis

All interviews will be audio recorded, transcribed verbatim and anonymised. Thematic analysis of parental attitudes and views around their use of social media for phenotypic data and novel approaches(13) will be used with NVivo to aid data management. Interview transcripts will be read and reread individually, from which an initial coding framework will be developed. Twenty percent of transcripts will be double coded. Team members will meet to discuss the preliminary coding framework and themes to ensure that the emerging analysis is trustworthy and credible. This framework will be added to and refined, with coded material regrouped as new data from subsequent interviews are gathered.

### Patient and public involvement

Discussions with parents of children with genetic syndromes and who are currently using services, colleagues providing health services to this patient group and feedback from public engagement events have all highlighted the need for more information to be available for families. Parents agreed there is a research gap, “when your child gets diagnosed there is so little information out there.” Another mother confirmed, “We are all used to going to medical appointments and witnessing a bit of ‘scratching of heads and then Googling!” They agreed clearer clinical guidance would be helpful, “often the doctors don’t know whether what is going on is just part of the genetics or something that still needs to be looked into.” The parents all felt that they wanted to be included in this research and the methods were acceptable, “this is answering questions, and most can be done from home.” Regarding recruitment, “we know that with rare things research is important and there aren’t many opportunities to get involved so we are keen to be involved in as much as we can.” One mother is lead administrator for a syndrome Facebook group of 120 patients and their families. She said that, “on the whole we’re just a big ‘family’ who share quality information and support each other – much of which isn’t available anywhere else…. “I think this is an amazing initiative, there is SO much to learn from groups like ours and we’re very happy to help with your project!” Another mother, (member of international and UK syndrome support group) remarked, “We know so much….so use us!”

Six parents have agreed to form a patient advisory group (PAG) meeting twice a year for the duration of the study. The PAG also includes a young adult who has a genetic syndrome and who is supported in attending the PAG meetings by her mother. The PAG is co-chaired by a parent member and there is also representation from Unique, a rare genetic disorders support charity. The PAG members will assist in the development of patient facing materials, advise on recruitment issues, inform the development of the topic guide for the qualitative interviews and discuss and help interpret the results. They will be offered a study-specific induction pack and relevant study information. Training workshops run by People and Research West of England will also be offered to them. PAG members will have their travel expenses and meeting time reimbursed with payment or vouchers. Our findings will be presented in lay terms at a PAG meeting, and they will advise us on routes for dissemination to patient groups.

### Ethics, monitoring and dissemination

This manuscript is based on Protocol V.7.0 dated 06/02/2023. The study received East Midlands -Nottingham Research Ethics Committee (REC) approval on 15 December 2022 and Health Research Authority approval on 9 February 2023. The study will be conducted in accordance with the protocol, NIHR Good Clinical Practice (14) and the principles of the Declaration of Helsinki. Any amendments of the protocol will be submitted to the REC for approval. On request, the study investigators and their institutions will permit study-related audits by the sponsor and relevant research ethics committee by providing direct access to source data. The University of Bristol holds the relevant insurance for this study and is the nominated sponsor for this study. A stakeholder group has been set up including clinical and laboratory geneticists and representatives from Unique and the Genomic Alliance. The stakeholder group provides advice to the investigators on all aspects of the study including aspects of safety and monitoring of serious adverse events. The DECIPHER project was given a favourable NRES REC opinion by Cambridge South (previously Cambridgeshire 4 REC), REC reference 04/MRE05/50, in November 2004. DECIPHER submits annual progress reports to ensure this favourable opinion applies for the duration of the research.

### Dissemination

A lay summary of the proposed study is available on the National Institute for Health Research website. Results of this study will be disseminated to those involved and publicly available through open access publication in peer-reviewed journals and presented at relevant conferences and research meetings. The PPI groups will contribute to the dissemination plan and assist in the production of the lay summaries.

Legends:

Supplementary figure 1: Eligible gene list

## Supporting information

Supplementary figure 1: Eligible gene list

## Data Availability

As this is a protocol paper there is no data requiring repository.

## Acknowledgements

The GenROC study was originally conceptualized with significant mentorship and guidance from Professor Esther Crawley who has now retired. We thank her for her significant contribution to the concept of the GenROC study.

The GenROC study will only be possible with involvement from multiple individuals across the UK. This forms the GenROC Consortium. A full list of contributors to the GenROC Consortium is available at https://www.bristol.ac.uk/academic-child-health/research/research/genetics/genroc-study/

## Author contributions

KJL led concept and design of protocol, led on ethics approval process, led PPI work, drafting and revision of manuscript. AW input to design, approvals and manuscript. PSB input to statistical analysis plans and manuscript. IN input to protocol design and manuscript. JP input to protocol design and manuscript. SLW input to PPI, recruitment pathway, dissemination plan and manuscript. JF input to protocol and manuscript. HVF input to protocol design and manuscript. JCI input to protocol design and manuscript.

## Data statement

As this is a protocol paper there is no data requiring repository.

