## Supplementary figure 1: Eligible gene list for "Improving the care of children with GENetic Rare disease: Observational Cohort study (GenROC): a study protocol"

| | | | | |
|---|---|---|---|---|
| ACTB | CSNK2A1 | ITPR1 | NSD2 | SMC1A |
| ACTL6B | CTCF | KANSL1 | OPHN1 | SMC3 |
| ADNP | CTNNB1 | KAT6A | PACS1 | SON |
| AHDC1 | CUL4B | KAT6B | PHIP | SOX5 |
| ANKRD11 | DDX3X | KCNQ2 | PIGN | SPTAN1 |
| ASH1L | DEAF1 | KDM5B | POGZ | SRCAP |
| ASXL3 | DPF2 | KDM5C | PPP2R5D | STXBP1 |
| ATP1A3 | DYNC1H1 | KIF1A | PRMT7 | SYNGAP1 |
| ATRX | DYRK1A | KMT2A | PUF60 | TAF1 |
| AUTS2 | EBF3 | KMT2C | PURA | TBL1XR1 |
| BCL11A | EEF1A2 | KMT5B | RAI1 | TCF20 |
| BPTF | EFTUD2 | LZTR1 | RERE | TCF4 |
| BRPF1 | ERF | MAGEL2 | RPS6KA3 | TLK2 |
| BRWD3 | FBX011 | MECP2 | SATB2 | TRAPPC9 |
| CACNA1A | FOXG1 | MED12 | SCN1A | TRIO |
| CAMTA1 | FOXP1 | MED13 | SCN1B | TRIP12 |
| CASK | GATAD2B | MED13L | SCN2A | TUBA1A |
| CDK13 | GLI2 | MEF2C | SCN8A | USP9X |
| CHD2 | GRIK2 | MYT1L | SETD1A | VPS13B |
| CHD3 | GRIN1 | NAA10 | SETD5 | WAC |
| CHD4 | GRIN2A | NAA15 | SHANK2 | WDR45 |
| CHD7 | GRIN2B | NALCN | SHANK3 | WDR62 |
| CHD8 | HECW2 | NEXMIF | SIN3A | WDR73 |
| CLTC | HNRNPU | NFIX | SLC6A1 | ZBTB20 |
| CNOT3 | HUWE1 | NRXN1 | SLC6A8 | ZMYND11 |
| CNTNAP1 | IQSEC2 | | SLC9A6 | |
